# Effectiveness of a school-based high-intensity interval training intervention in adolescents: study protocol of the *PRO-HIIT* cluster randomised controlled trial

**DOI:** 10.1101/2024.06.28.24309651

**Authors:** Yong Liu, Alan R. Barker, Anna-Lynne R. Adlam, Minghui Li, Stephanie L. Duncombe, Andrew O. Agbaje, Yaodong Gu, Huiyu Zhou, Craig A. Williams

**Affiliations:** Children’s Health and Exercise Research Centre, Public Health and Sports Sciences, Faculty of Health and Life Sciences, University of Exeter, Exeter, United Kingdom; Psychology, Faculty of Health and Life Sciences, University of Exeter, Exeter, United Kingdom; Faculty of Sports Science, Ningbo University, Ningbo, China; School of Public Health, The University of Queensland, Herston, QLD, Australia; Institute of Public Health and Clinical Nutrition, School of Medicine, University of Eastern Finland, Kuopio Campus, Kuopio, Finland

**Author notes:** corresponding author: Craig A. Williams.

## Abstract

**Introduction:** High-intensity interval training (HIIT) is an effective strategy for improving a variety of health and fitness outcomes within the school settings. Incorporating HIIT into existing physical activity opportunities appears practically feasible, yet the process evaluation and effectiveness of this strategy needs to be further evaluated. Therefore, a *PRO-HIIT* intervention will be conducted to evaluate the effectiveness of a 12-week school-based HIIT intervention on cardiorespiratory fitness, physical activity, body composition, muscular strength, bone health, cognitive function, wellbeing and academic performance among 12-13-year-olds.

**Methods and analysis:** Eight classes of year 7 students (12-13-year-olds) from a secondary school in Ningbo, China, will be recruited and randomly allocated into an intervention or control group. While the control group maintains their usual activities, a 6 to 10-minute HIIT session will be embedded in the physical education or physical activity lessons five days a week for 12 weeks for the intervention group. Training workshops will be conducted for participants, teachers, and research staff for facilitating the intervention. Outcome data will be collected at three time points: pre- and post-intervention, and two months (summer holiday) upon completion of the intervention. Linear mixed models will be used to analyse the impact of groups (intervention and control), timepoints (pre-, post- and two-month after intervention) and group by time interactions. The implementation process of the intervention will be evaluated with the guidance of MRC process evaluation framework.

**Ethics and dissemination:** Ethics approval is obtained from the Ningbo University Ethics Committee (TY2024002) and the Public Health and Sport Sciences ethics committee, University of Exeter (5713479). Results from *PRO-*HIIT study will be disseminated via peer-reviewed journals, scientific conferences as well as local education system. The study protocol has been retrospectively registered on ClinicalTrials.gov Protocol Registration and Results System (NCT06374732), https://clinicaltrials.gov/study/NCT06374732.

## Introduction

Physical activity (PA) is well documented for its role to promote physical fitness and mental wellbeing in children and adolescents (1–4). However, less than 20% of adolescents meet the World Health Organization guideline for engaging in an average of 60 minutes of daily moderate to vigorous PA across the week (3). The most cited barriers for adhering to the recommendations include time constraints, lack of motivation and inadequate facilities (5, 6). Notably, engaging in vigorous PA is of salient health significance for children and adolescents. Vigorous PA has been independently associated with lower levels of cardiometabolic risk factors, increased cardiorespiratory fitness (CRF) and improved bone health in this cohort (7, 8). With time constraints for PA participation among young population, prioritising vigorous PA by employing high-intensity interval training (HIIT) might be a viable option for health promotion (9). HIIT, featuring short bursts of intense exercises interspersed with active recoveries or rests, has emerged as a time-efficient and effective exercise strategy for children and adolescents (5, 10).

Schools act as the ideal settings for PA programmes because of the abundant resources available within the education system, such as staff, space and facilities, different timing options (e.g., breaks and classes) and broad reach of children and adolescents (1, 11). Time spent in schools account for a significant proportion of children’s waking hours, hence these institutions have the potential to counteract the global issues related to physical inactivity (12) and health inequalities (13). In recent years, there has been a surge in integrating HIIT into school settings. Review-based evidence indicates that school-based HIIT interventions effectively enhance body composition (14–16), CRF (15, 16) and muscular health (17). However, the impact on cognitive function and academic performance remains uncertain (15) and evidence regarding its health benefits for mental well-being (15) and bone health (18) is limited.

Despite the advantages of school settings, integration of PA programmes into schools is challenging. This is mainly because of the additional workload imposed on already overworked schoolteachers and the potential diversion of students’ valuable time away from academic study (19). One practical solution to address this challenge is to incorporate PA interventions during dedicated curriculum time for PA, such as during physical education (PE) lessons. Lubans et al. (9) proposed that for school-based HIIT to be scalable, it should be integrated into existing PE or sport training sessions. Indeed, a recent review highlighted that 57.1% (n = 24) of the 42 identified school-based HIIT interventions were conducted during PE lessons (15). Given that less than 50 % of a typical PE lesson time is spent in moderate-to-vigorous PA in secondary school (20), HIIT targeting PE lessons might potentially enhance the quality of PE and health condition of children and adolescents (21).

Recent research has highlighted the risk of fitness loss or stagnation during prolonged and unstructured days (defined as weekends or holidays when obesogenic behaviours are prevalent due to lack of compulsory PA opportunities, restriction on caloric intake, limitation on screen time, and regulated sleep schedules), a phenomenon known as the “Structured Days Hypothesis (SDH)” (22). Supporting the SDH, a survey conducted in Australia demonstrated that children spent more time watching TV or playing videogames and engaged in less PA, leading to a reduced daily energy expenditure during unstructured holidays (23). Martin et al. (24) showcased the efficacy of a 7-week school-based HIIT programme in mitigating potential CRF loss among Scottish adolescents during summer vacation. However, the impact on other health-related factors remains underexplored and their study is limited by a lack of implementation details.

One useful approach to improve the intervention reporting is through conducting a process evaluation. Process evaluation delves into implementation details, mechanisms of impact and contextual factors, offering a comprehensive understanding regarding the intervention effectiveness and underlying causality (25). It runs parallel to the outcome assessment, which contributes to future scaling up and dissemination (25, 26). Nonetheless, only a limited number of school-based HIIT interventions have included process evaluations, either as sections within intervention outcome papers (27, 28) or as standalone pieces (29–31). Among these studies, only two interventions employed a process evaluation framework (29, 31). Conducting process evaluation without proper guidance may pose challenges, potentially leading to incomplete reporting and biased results and interpretations (32).

Given the above stated research gaps, we aim to conduct a school-based HIIT intervention named *PRO-HIIT.* The *PRO-HIIT* is a health promotion initiative designed to deliver 6-10 minutes of HIIT into the daily routines of Chinese adolescents, with a focus on settings of existing PA opportunities. Specifically, while the control group will take the usual PE (n = 3) and PA lessons (n = 2) every week, a 6-10-minute of HIIT will be embedded at the beginning of these lessons five times per week for twelve weeks for the intervention group. The aims of the *PRO-HIIT* are to:

1. evaluate the effectiveness of the *PRO-HIIT* on CRF (primary outcome), PA, body composition, muscular strength, bone health, executive function, wellbeing, enjoyment, motivation, affect, self-efficacy and academic performance among 12-13-year-old adolescents;
2. to examine the changes of these outcomes following a two-month unstructured summer holiday after the *PRO-HIIT* study is completed;
3. to evaluate the implementation process of the *PRO-HIIT* through a process evaluation.

## Materials and methods

### Study design

The study is a multi-centre collaborative work conducted by University of Exeter and Ningbo University. Consolidated Standards of Reporting Trials (33) and Template for Intervention Description and Replication (34) checklists will be adopted for guidance and reporting. The *PRO-HIIT* study will employ a two-arm cluster-randomised controlled trial design, with an intervention group and a treat as usual control group. Clusters in this study are eight classes of secondary school students, located in Ningbo City, China. The intervention will be delivered five times per week, commencing at the beginning of the three PE and two PA lessons. The PA lessons serve as a complementary opportunity for students to engage in exercises of their choice on days when PE lessons are not scheduled. Assessments will occur at three time points: baseline (T1), immediate post-intervention (T2) and two-month post-intervention (T3), with T3 aligning with the initial two weeks of a new school term following a two-month unstructured summer holiday. Fig 1 presents an overview of the schedule of enrolment, interventions, and assessments.

**Fig 1.**
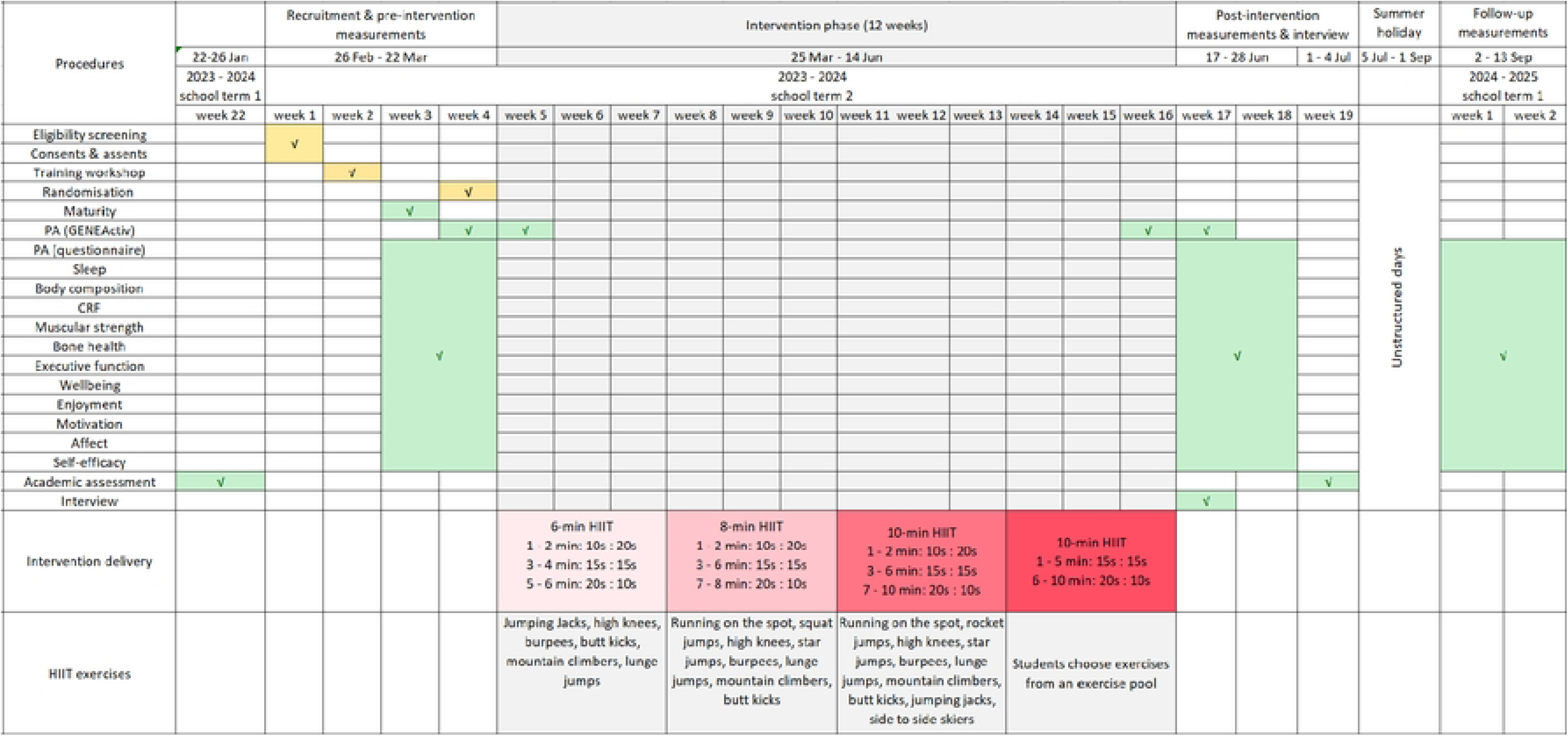
The overview of the schedule of enrolment, interventions, and assessments for *PRO-HIIT*. HIIT, high-intensity interval training; PA, physical activity; yellow, preparation for the intervention; green, measurements and data collection; red, progressively increased exercise intensity and volume; grey, intervention period.

### Sample size estimation

A sample size estimation was calculated based on CRF using the 20-m shuttle run test (20m-SRT). Previous studies reported 9 laps of improvement, with effect size of *d* = 0.31 (35) and *d* = 0.69 (36), achieved through 12 or 14 weeks of resistance-based HIIT, respectively. In the current study, we utilised a conservative effect size of *d* = 0.31 for power calculation. Therefore, based on a G*Power (Version 3.1) calculation (37) and using two groups with 80 % power at an α of 0.05, it is estimated to recruit 165 participants in each group. With an average enrolment of 50 students in secondary school classes in Ningbo and a conservative 20 % of dropout rate (38), it is deemed that 8 classes (approximately 50 participants per class) would provide sufficient statistical power for analysis. Given the typical scale of a Chinese secondary school (10 - 15 classes per grade), the 8 classes will be recruited from one secondary school.

### Recruitment and allocation

The recruitment started from 26^th^ February and ended on 1^st^ March 2024. An invitation letter will be sent to nearby secondary schools at Ningbo University. When school(s) express their interest, we will initiate contact to elucidate our participation requirements. Upon agreement from the school(s), we will extend invitations to head teachers and PE teachers of year 7, who will then present the study to students for recruitment. To be eligible for participation, students are required to submit signed assent forms along with signed consent forms from their parents/guardians. Students with health or medical conditions that would restrict their ability to engage in vigorous PA will be excluded from the study. The screening process will involve reviewing the medical examination reports submitted by the participants’ parents or guardians. Classes will be randomly allocated to either *PRO-HIIT* group or control group, via a computer-based random number generator by an independent researcher. The randomisation and allocation will take place after baseline assessment. The randomisation will be stratified by PE teacher, wherein each teacher’s classes will be randomly assigned to either the intervention or control group.

### Intervention delivery and exercise design

The intervention will be delivered five times a week for twelve weeks (school term 2, week 5 – week 16, March – June 2024) at the beginning of the three PE lessons and two PA lessons. Throughout the intervention periods, PE teachers will coordinate and supervise the HIIT intervention, while the delivery will be accomplished by two student peer-coaches, selected by the PE teacher within each intervention class.

To ensure quality and effective implementation, the leading researcher will collaborate with the PE teachers and peer-coaches for delivering the intervention during the initial two weeks. Researchers will have a school visiting once per month to provide on-going support and guidance. Additionally, a training logbook at each intervention class will be maintained by the PE teachers to document aspects, such as attendance, dose delivered and received, and adverse events. These records will be sent to researchers on a weekly basis to ensure prompt feedback and communication. In cases where HIIT sessions are cancelled due to inevitable factors such as large school events or severe weather, participants will be encouraged to complete these sessions during alternative times (e.g., breaks). These additional sessions will be supervised by peer-coaches and will be recorded in the training logbook. The control group will take their regular PE and PA lessons as usual.

The HIIT sessions consist of body-weight resistance exercises (e.g., high knees, jumping jacks, burpees), selected based on relevant literature (32, 39). The session length will be progressively extended from 6 to 10 minutes over the 12 weeks, accounting for fitness adaptations. Within each session, the work-to-rest ratio will increase from 10s: 20s, to 15s: 15s, until 20s: 10s as the exercise progresses. Furthermore, participants will perform one exercise (e.g., jumping jacks) twice in a minute to avoid monotony while preventing frequent exercise changes, thereby 6, 8 and 10 different exercises will be completed as the intervention duration increases. Moreover, flexibility will be allowed for each HIIT session, enabling adaptations whenever necessary (e.g., higher/lower exercise intensity). The adaptations made will be recorded on the training logbook. The details regarding the exercises are presented in Fig 1.

To facilitate the implementation process, several strategies will be employed, including:

1. training workshops for participants, peer-coaches, teachers and research staff (Table 1);
2. providing participants with opportunities to choose music and exercises (from an exercise pool over the final 3 weeks of the intervention);
3. the opportunity to win a prize upon completion of the intervention for all participants.

**Table 1.** Details of training workshops for participants, peer-coaches, teachers and research staff.

| Subjects | Timing | Content |
| --- | --- | --- |
| Participants (CON) | Two PE lessons (40 minutes) | 1) Introduction of the programme; 2) outcome measurements; 3) familiarisation of executive function tasks; 4) maintain daily life. |
| Participants (INT) | Two PE lessons (80 minutes) | 1) Introduction of the programme; 2) familiarisation of equipment (e.g., HR monitor), executive function tasks and HIIT exercises; 3) outcome measurements. |
| Peer-coaches | One PE lessons (40 minutes) | 1) HIIT performing and leading; 2) HIIT rescheduling and regulation. |
| PE teachers | 60 minutes | 1) Introduction of the programme; 2) familiarisation of equipment (e.g., HR monitor); 3) HIIT exercises; 4) training logbook; 5) intervention delivery. |
| Research staff | 60 minutes | 1) Outcome measurements; 2) data collection principles (e.g., SAAFE). |

CON, control group; INT, intervention group; PE, physical education; HR, heart rate; HIIT, high-intensity interval training; SAAFE, The Supportive, Active, Autonomous, Fair, Enjoyable principles.

### Theoretical frameworks

The present study draws upon guidance from two frameworks: process evaluation of complex interventions: Medical Research Council (MRC) guidance (25) and the Supportive, Active, Autonomous, Fair and Enjoyable (SAAFE) principles (40).

### Process evaluation

A comprehensive process evaluation will be conducted for the *PRO-HIIT* intervention, which will be guided by the MRC process evaluation framework (25). The framework contains three domains, including implementation (i.e., fidelity, reach, recruitment and retention, dose delivered and adaptation), mechanisms of impact (i.e., mediators, dose received, unintended consequences and response) and context (i.e., barriers, facilitators, and contamination). The MRC framework, in conjunction with insights from a recent school-based HIIT review by Liu et al. (26), will guide the adaptation of process evaluation measures tailored specifically to *PRO-HIIT*, as detailed in Table 2.

**Table 2.**
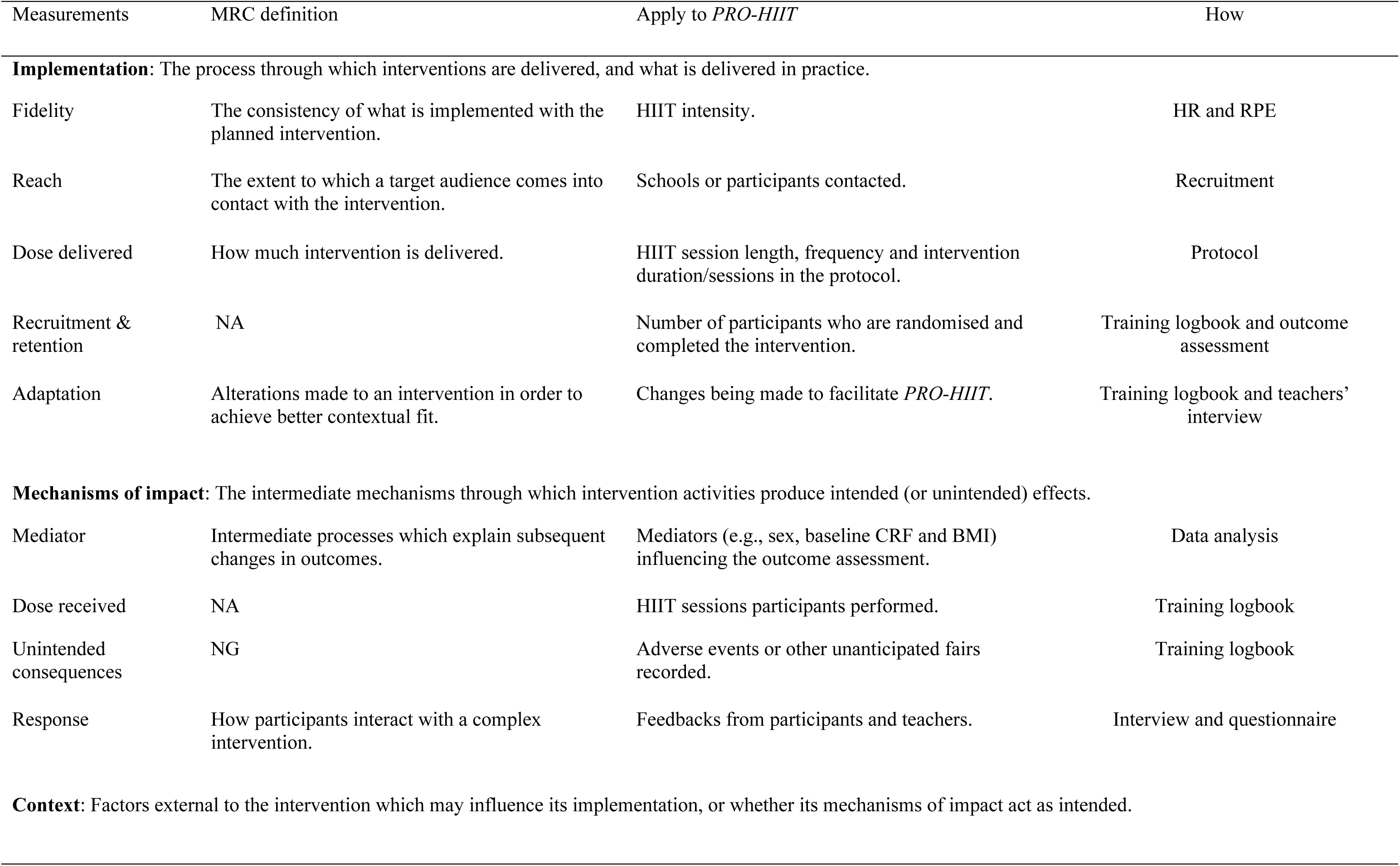
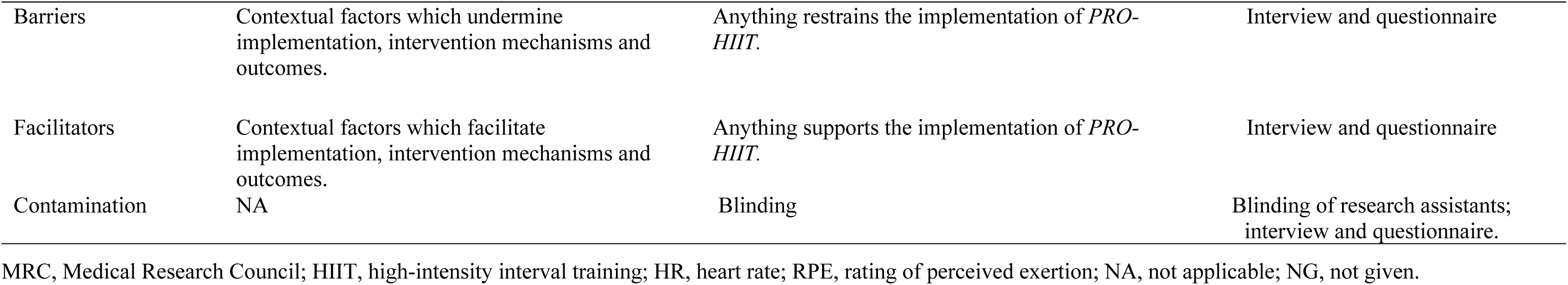
Medical Research Council definition for process evaluation and its application to *PRO-HIIT*.

It is important to highlight that while all the intervention classes will be encouraged to exercise with “all out” efforts during the HIIT sessions, the intensity will be monitored in only two randomly selected classes on one occasion per week (during one of their PE lessons) to minimise extra burden for PE teachers. The intensity in these two classes will be measured using either heart rate (HR) or rating of perceived exertion (RPE) by Polar Verity Sense and OMNI Perceived Exertion Scale for resistance exercise (41), respectively. Of note, due to resource constraints, only 10 Polar Verity Sense monitors will be utilised and rotated weekly among participants in the HR monitoring class. However, all participants will be required to report their peak RPE during the HIIT session once per week in the RPE monitoring class.

### SAAFE principles

SAAFE principles offer a structured framework designed to inform the design, delivery, and evaluation of PA interventions (40). SAAFE principles provide essential guidelines and practical strategies to enable practitioners to organise their interventions in a way that not only maximises PA participation but also fosters positive affective, cognitive, motivational, and movement skill outcomes. The principles and strategies will be incorporated into the exercise design, intervention delivery and outcome assessment processes of the *PRO-HIIT* intervention (Table 3). These principles will be applied or achieved through the process of exercise design, training workshops, HIIT session delivery and outcome assessments.

**Table 3.** Definition and application of SAAFE in *PRO-HIIT* intervention.

| Principles | Definition | Apply to *PRO-HIIT* | How |
| --- | --- | --- | --- |
| Supportive | Intervention is designed to facilitate a supportive environment | Encourage praise of students' effort and improvement during HIIT sessions and outcome evaluation process. | Researchers and teachers' training workshop. |
|  |  | Encourage mutual support when performing HIIT and outcome assessments. | Students' training workshop. |
|  |  | Demonstrate empathy toward students when they feel frustrated or challenged. | Researchers and teachers' training workshop. |
| Active | Sessions are highly active | Sessions are designed without any instruction time. | Exercise design. |
|  |  | Exercises are performed at high intensity. | Encourage exercise with “all out” efforts. |
| Autonomous | Sessions involve elements of choice | Right to play any music they like. | Students' training workshop. |
|  |  | Self-organisation. | Peer coaches to lead the HIIT sessions. |
|  |  | Right to choose exercises from an exercise pool (final 3 weeks). | Every Friday prior to the week the exercise will be performed. |
|  |  | Perform the missed HIIT sessions themselves during breaks. | Under the supervision of peer-coaches and record on training logbook. |
|  |  | Minimize controlling language. | Researchers and teachers' training workshop. |
| Fair | Intervention provides all students with opportunities to experience success | Encourage self-comparison rather than peer-comparison. | Students' training workshop. |
|  |  | Provide personalised care for individuals with special needs (e.g., participants with lower fitness levels). | Simplify exercises or lower exercise intensity (PE teacher' discretion). |
| Enjoyable | Intervention is designed to be enjoyable and engaging for all students | Provide different HIIT workouts. | From existing literature. |
|  |  | Provide challenging HIIT sessions. | Progressively increase the exercise duration and intervals within and across sessions. |
|  |  | Play music while exercising. | Peer-coaches. |

HIIT, high-intensity interval training.

### Outcome measurements

Outcome measurements will be conducted in the school premises by trained research assistants who will remain blinded to group allocation throughout all assessment time-points. All measurements will be introduced and explained during the training workshop and will be described again prior to the commencement of the measurements to ensure clarity. The measurements will be conducted during their regular PE lessons. Details of the outcome measures are summarised in Table 4.

**Table 4.** Outcome variables, tools and timing for outcome measurements.

| Outcomes | Tools | Timing | How and where (all participants, measured at class level) |
| --- | --- | --- | --- |
| PA | GENEActiv | One week before and after intervention, first and last week of intervention. | A subsample of 60 participants (30 in each group, 30 males) will be randomly selected to wear the GENEActive accelerometer for 14 days consecutively for twice, covering the total 4 weeks. |
| CRF | 20m shuttle run | T1, T2, T3 | Multiple measurement stations will be held, and participants will be split into groups to ensure efficient measurements. Two PE lessons is estimated to be sufficient. All these measurements will be conducted in sports hall. |
| Body composition | TANITA | T1, T2, T3 |  |
| Muscular strength | Handgrip, standing long jump | T1, T2, T3 |  |
| Bone health | GE Achilles | T1, T2, T3 |  |
| Executive function | Flanker tasks, visual 2-back tasks, colour-shape switch tasks | T1, T2, T3 | Perform the executive function tasks online in a school computer room in one PE lesson time (40 minutes). |
| Wellbeing | SDQ | T1, T2, T3 | Finish together using one PE lesson time (40 minutes) in a school computer room with the instruction of research staff. |
| Enjoyment | PACES | T1, T2, T3 |  |
| Motivation | BREQ-2 | T1, T2, T3 |  |
| Affect | PANAS-SF | T1, T2, T3 |  |
| Self-efficacy | PPAS | T1, T2, T3 |  |
| Academic performance | End-of-term academic examinations | By the end of term 1 and T2 | Obtained directly from head teacher. |

PA, physical activity; CRF, cardiorespiratory fitness; SDQ, Strength and Difficulty Questionnaire; PACES, Physical Activity Enjoyment Scale; BREQ-2, Exercise Regulations Questionnaire; PANAS-SF, Positive and Negative Affect Schedule Short Form; PPAS, Perceived Physical Ability Scale; T1, pre-intervention; T2, immediately post-intervention; T3, 2-month follow-up.

### Primary outcome

#### Cardiorespiratory fitness

CRF will be assessed using the 20m-SRT, a well-established field test for evaluating CRF (42). The 20m-SRT requires participants to run back and forth between two lines positioned 20 meters apart. Participants must reach the other end before a designated beep sounds. The beep is set to allow the participants to start at 8.5 km/hour with increases in speed of 0.5 km/hour denoted by a triple beep. The test concludes if a participant fails to complete two consecutive shuttles or volitionally discontinues. Performance on the 20m-SRT will be reported as number of laps completed. The test will be administered by the same group of research assistants at the same location, time of the day and with consistent levels of verbal encouragement across all measurement timepoints to avoid biased results.

### Secondary outcomes

#### Physical activity

For a subset of randomly selected (stratified by sex and group) participants (n = 60), PA will be objectively assessed using GENEActiv wrist-worn accelerometers (Model GAT04, Activinsights Ltd, Cambridgeshire, England) over a span of four weeks, comprising one week before and after the intervention as well as the initial and concluding weeks of the intervention. Participants will be encouraged to wear the device 24 hours/day, without taking off even when bathing or sleeping. GENEActive accelerometers have demonstrated acceptable reliability and validity for PA monitoring in adolescents (43). The cut-points employed to categorise sedentary, light, moderate and vigorous PA are < 6, 6 – 21, 22 – 56, and > 56 g s, respectively (43).

#### Body composition

Height and waist circumference will be measured with a portable stadiometer and a tape, respectively. Subsequently, waist-to-height ratio will be calculated (44). Body mass and body fat percentage will be determined with a Tanita device (Tanita Corp., Tokyo, Japan).

#### Muscular strength

Upper and lower body strength will be assessed using hand grip and standing long jump, respectively. A digital dynamometer with an adjustable grip, with participants standing and elbow in 90-degree flexion, will be employed for measuring the upper body strength in kilogrammes (45). The test will be performed once on both hands and the highest record will be reported. Standing long jump will be measured with a standing long jump mat. Participants stand behind the start line with their feet apart and are allowed to swing their arms quickly to jump as far as possible. Each participant will have three attempts, with the best one recorded.

#### Bone health

A heel ultrasound test will be performed via a GE Achilles heel ultrasound machine (GE Medical Systems Lunar, USA). Participants will be seated with one foot on the foot plate, and alcohol will be applied to ensure proper membrane contact. A transducer on one side of the heel will convert an electrical signal into a sound wave, which will pass through the heel to the other side and be received and analysed by another transducer. The speed of sound (SOS, in m/s) and broadband ultrasound attenuation (BUA, in dB/MHz) will be measured and used to calculate the stiffness index (SI) with the equation: SI = (0.67 × BUA + 0.28 × SOS) − 420 (46). The test will be performed on both feet and an average score will be recorded.

#### Executive function

Participants’ executive function will be assessed on aspects of working memory, inhibition and cognitive flexibility (47). Three tasks will be utilised, including flanker task, visual 2-back task and colour-shape switch task. The tasks are adapted from studies conducted by Wassenaar and colleague (48) and will be programmed on the Gorilla platform (49). The order of the three tasks will be randomised at individual level and will be performed collectively within the school computer room on a class-unit basis, with the presence of researchers to provide clarification if needed. One week prior to the intervention, a training workshop will be conducted to acquaint participants with the executive function tasks. A ten-minute presentation will elucidate the task procedures by research staff, and participants will engage in hands-on practise for each task. Any questions and inquiries will be addressed within the workshop to ensure clarity and understanding. Details for the three tasks are provided in the additional file 1.

#### Wellbeing

The Chinese version of the Strengths and Difficulties Questionnaire (SDQ) will be used to assess the psychological distress of participants (50). The questionnaire will be administered online via the Gorilla platform following the completion of executive function tasks. The SDQ comprises 25 personality items, rated on a 3-point scale (i.e., *‘not true’=0, ‘somewhat true’=1* and *‘certainly true’=2*), and is composed of five subscales, each consisting of 5 items. These subscales include emotional symptoms, conduct problems, hyperactivity/inattention, peer relationship problems and prosocial behaviour. The total difficulties score ranges from 0 – 40, where a score ≥ 17 is considered as high difficulties (51).

#### Enjoyment

Enjoyment of PA will be measured using the Physical Activity Enjoyment Scale (PACES) (52), which is validated among adolescents (53). The scale commences with a prompt “when I am active” followed by 16 phrases that participants will rank on a 5-point scale, ranging from 1 (Disagree a lot) to 5 (Agree a lot). The enjoyment score ranges from 16 – 80, with a higher score representing higher level of PA enjoyment (53).

#### Motivation

Motivation to autonomously engage in PA will be assessed using a modified Behaviour Regulation in Exercise Questionnaire (BREQ-2), which is an 19-item validated questionnaire (54). The scale comprised of 5 subscales, including intrinsic, identified, introjected, external and amotivation. Each item was rated on a 5-point Likert scale ranging from 0 (“not true for me”) to 4 (“very true for me”). The mean of the 5 subscales will be calculated to reflect the extent of each motivation type separately. A Relative Autonomy Index will be adopted by weighting (intrinsic * 3, identified * 2, introjected * −1, external * −2 and amotivation * −3) the subscales and summing the weighted scores (55). The Relative Autonomy Index ranges from −24 to 20 and higher positive scores indicate more autonomous motivation.

#### Affect

Affect will be assessed via a Chinese version of the International Positive and Negative Affect Schedule Short Form (PANAS-SF) (56). This 9-item questionnaire utilises a 5-point Likert scale, ranging from 1 (Not at all) to 5 (Extremely). Comprising 5 items related to positive affect and 4 items pertaining to negative affect, this instrument is considered reliable for its implementation in Chinese adolescents (56). The positive and negative affect scores will be summed and reported separately, with higher positive score indicates more positive affect and lower negative score indicates less negative affect.

#### Self-efficacy

A 6-item validated Perceived Physical Ability Scale (PPAS) will be utilised to evaluate the PA-related self-efficacy (57). In each item, four statements related to capabilities for doing exercises will be given and participants will be required to choose the one that best representing their personal feelings. The total score ranges from 1 to 24, with higher scores indicate a higher self-perception of physical ability and vice versa.

#### Enjoyment and satisfaction of HIIT

Enjoyment and satisfaction of the HIIT workout will be evaluated using a 2-item 5-point Likert scale, with the prompt: “I enjoyed/liked the HIIT workouts” and “I will continue to perform/use the HIIT workouts” between 1 = strongly disagree and 5 = strongly agree. This will only be assessed post-intervention among participants in the intervention group (T2).

#### Academic performance

Academic performance will be evaluated by utilising the school’s end-of-term academic examinations, which comprehensively assess all the subjects. Mathematics, language learning (main subjects in Chinese secondary school) and a composite score for all subjects will be utilised to discern variations in academic performance between the intervention and control groups. Therefore, academic performance will not be assessed at the follow-up stage.

#### Interview

Once the intervention completed, semi-structured interviews will be conducted with participants and PE teachers, separately. The two peer coaches and two participants (randomly selected) from each intervention classes (n = 16) will be invited to the participants’ interview, while all the PE teachers involved in the study will take part in the teachers’ interview. Pre-determined open-ended questions will be asked during the interview, including feedback on enjoyment/usefulness of the *PRO-HIIT* intervention, continued use of the HIIT exercises and the perceived barriers and facilitators for doing/delivering the HIIT exercises. The template of the interview questions is provided in the additional file 2.

#### Confounding variables

An estimate of the age of peak height velocity will be used to assess the somatic maturation of participants (58). Participants’ PA and sleep at all time-points will be assessed by a Chinese version of the International Physical Activity Questionnaire, short form (IPAQ-SF) (59) and a validated Chinese version of the Pittsburgh Sleep Quality Index (PSQI) (60), respectively.

#### Statistical analyses

Data entry will be completed by one researcher with a random sample of at least 10 % of entries cross-checked by a second researcher for accuracy. Prior to analysis, thorough checks for outliers and errors will be conducted using range and boxplot methods. Additionally, assessments for normality, homogeneity of variances and sphericity will be conducted as needed to verify assumptions. The baseline data for intervention and control groups will be presented and compared at individual level using independent sample t-test. An intention-to-treat approach will be adopted to evaluate the effects on outcome variables to avoid bias in exploring the impact of the intervention. Linear mixed-effect models will be used, with random effects, to analyse the impact of groups (intervention and control), timepoints (pre-, post-intervention and follow-up) and group x time interactions. Statistical analyses will be adjusted for the clustering effects at class level. Per protocol sensitivity analysis will be undertaken at the class level. Considering for disruptions such as school holidays, exams, severe weather, a minimum of 30 sessions is considered achievable over the 12-week period. Other sensitivity analyses, such as complete-case analysis, will be conducted where appropriate. Moderators, including sex (male, female), baseline overweight/obese (yes, no) and baseline CRF (healthy vs. needs to improve), will be examined with linear mixed models. Where appropriate, subgroup analyses will be conducted for the significant group-by-moderator interactions. PA, sleep and maturity will be included in the model to eliminate confounding effects. All data analyses will be conducted via IBM SPSS Statistics for Windows (SPSS 28.0; IBM Corporation, Armonk, NY, USA), with an alpha level of 0.05.

#### Patient and public involvement

Important input and feedback were sought from school leaders, teachers and students in the secondary schools located in Ningbo to inform and refine the study design of the *PRO-HIIT* intervention.

#### Ethics and dissemination

The *PRO-HIIT* study is approved by the Clinical Research Ethics Committee, Ningbo University, China (TY2024002) and Department of Public Health and Sport Sciences ethics committee, University of Exeter (5713479). Appropriate checks and training will be completed for all the researchers before initiating the study to ensure the safety of participants. The school principals and involved teachers will provide consent. Participants and parents/guardians will provide consent and assent forms, respectively, to be included in the study. Data will be de-identified with a unique participant code and stored in a password-protected computer. The study protocol has been registered on ClinicalTrials.gov Protocol Registration and Results System (NCT06374732). Results of *PRO-HIIT* will be disseminated through publications in peer-reviewed journals and conference presentations. Meanwhile, our findings will be reported to the local education system for their consideration, and we will encourage the schools to continue using the *PRO-HIIT* protocol during their PA-related classes.

## Discussion

This paper outlines the study protocol for *PRO-HIIT* intervention, which aims to investigate the effectiveness of a school-based HIIT intervention on CRF, PA, body composition, muscular strength, bone health, executive function, wellbeing, enjoyment, motivation, affect, self-efficacy and academic performance in school-aged adolescents. The *PRO-HIIT* intervention will be delivered five times per week at the beginning of the three PE and two PA lessons. The PA lessons serve as a complementary opportunity for students to engage in exercises of their choice on days when PE lessons are not scheduled. The majority of school-based HIIT interventions were administered 2 to 3 times per week (15). However, the understanding of the feasibility and effectiveness of HIIT performed five times per week is limited, with only two studies identified (61, 62). Moreau et al. reported that a daily 10-minute HIIT session over 6 weeks improved cognitive control and working memory in children aged 7 to 13 years (61). In addition, a 10-month of 5 x 12 minute/week interval running programme was found to be feasible in a primary school setting (62). While the study reported a positive effect on sprint performance, no effects were observed on CRF, BMI, muscular fitness, and bone health. Consequently, the feasibility and effectiveness of high-frequency HIIT interventions delivered among secondary school students needs to be further evaluated. The *PRO-HIIT* study aims not only examine the effects on commonly studied variables such as body composition, CRF, muscular strength and cognition, but also to explore its impact on bone health and academic performance among 13-year-old adolescents in secondary schools.

The early pubertal phase is recognised as the time when peak bone mass accrual begins. High-impact exercises, such as resistance-based HIIT, may enhance bone mass accumulation during these crucial developmental years. Yet, there is a dearth of research investigating the association between HIIT and bone health (18). Emerging evidence suggested that both acute and chronic HIIT leads to enhanced cognitive adaptations and brain health (63, 64). Additionally, a longitudinal study revealed that children with higher fitness levels exhibit greater hippocampal volume and superior memory performance compared to their less fit counterparts (65). Given that executive function is closely linked to academic performance (64, 66), a long-term HIIT intervention has the potential to enhance academic performance. Nevertheless, only one previous study has examined the effect of HIIT on academic performances (67). The researchers found that HIIT delivered twice per week for ten weeks significantly improved academic performance on mathematics and language in the intervention group as compared to the control group. It is worth noting that this study was conducted in primary schools, warranting further investigation in diverse educational settings.

Apart from the outcome measurements, the novelty of *PRO-HIIT* study lies in its attention to the implementation process, a component often overlooked in previous school-based HIIT interventions (15). The present study will scrutinise the intervention process with the guidance of the MRC process evaluation guidelines, thereby enhancing the understanding and facilitating the dissemination of the *PRO-HIIT* study. Another novelty of the *PRO-HIIT* study is its aim to determine the extent of potential fitness loss over the two-month summer holidays and how a school-based intervention may mitigate this decline, thereby contributing to the examination of the SDH. Furthermore, existing HIIT interventions in schools have predominantly taken place in western countries (15), the *PRO-HIIT* study will contribute valuable insights by conducting similar interventions within an Asian cultural context.

## Authors’ contributions

The study design was conceived by Yong Liu, Alan R. Barker and Craig A. Williams. Yong Liu, Minghui Li, Huiyu Zhou and Yaodong Gu is responsible for the implementation and data collection of the intervention. Anna-Lynne R. Adlam, Stephanie L. Duncombe and Andrew O. Agbaje contributed to the data analyses. Yong Liu drafted the manuscript, while all authors contributed to the revision of the manuscript and approved for submission.

## Data Availability

No datasets were generated or analysed during the current study. All relevant data from this study will be made available upon study completion.

https://clinicaltrials.gov/study/NCT06374732

## Acknowledgements

Many thanks to teachers and students in Jiao Chuan Shu Yuan for participating in the intervention.

## Data availability

All relevant data for this study will be made available upon study completion.

